# Comparative Effects of Weight Loss and Incretin-Based Therapies on Vascular Endothelial Function, Fibrinolysis, and Inflammation: A Randomized, Controlled Trial

**DOI:** 10.1101/2022.02.23.22271434

**Authors:** Mona Mashayekhi, Joshua A. Beckman, Hui Nian, Erica M. Garner, Dustin Mayfield, Patricia Wright, Sara E. Howard, Bradley Perkins, Dianna Olson, William E. Snyder, Jessica K. Devin, John R. Koethe, Jonathan D. Brown, Katherine N. Cahill, Chang Yu, Heidi Silver, Kevin Niswender, James M. Luther, Nancy J. Brown

**Affiliations:** Vanderbilt University Medical Center, Department of Medicine, Division of Diabetes, Endocrinology and Metabolism, Nashville, TN; Vanderbilt University Medical Center, Department of Medicine, Division of Cardiovascular Medicine, Nashville, TN; Vanderbilt University Medical Center, Department of Biostatistics, Nashville, TN; Vanderbilt University Medical Center, Department of Medicine, Division of Clinical Pharmacology, Nashville, TN; Alabama College of Osteopathic Medicine, Dothon, AL; Vanderbilt University Medical Center, Department of Medicine, Division of Gastroenterology, Nashville, TN; Vanderbilt University Medical Center, Department of Medicine, Division of Allergy, Pulmonary and Critical Care Medicine, Nashville, TN; UCHealth Endocrinology, Yampa Valley Medical Center, Steamboat Springs, CO; Veterans Affairs Tennessee Valley Healthcare System, Nashville, TN; Vanderbilt University Medical Center, Department of Medicine, Division of Infectious Diseases, Nashville, TN; Yale School of Medicine, New Haven, CT

**Keywords:** Cardiovascular disease, DPPIV Inhibitor, Flow-mediated dilation, GLP-1 Receptor Agonist, Monocyte chemoattractant protein-1, Obesity, Plasminogen activator inhibitor-1, Prediabetes

## Abstract

**Aims/Hypothesis:** GLP-1 receptor (GLP1R) agonists cause weight loss in obese individuals and decrease cardiovascular events in patients with type 2 diabetes. The objective of this study was to test the hypothesis that GLP1R agonists have beneficial effects on vascular endothelial function, fibrinolysis, and inflammation in obesity through weight loss-independent (GLP1R-dependent) mechanism(s).

**Methods:** We conducted a randomized, parallel-group, three-arm double-blinded controlled trial at a single center in the United States. Eligibility criteria were as follows: age 18-65 years, body mass index ≥ 30kg/m^2^, and pre-diabetes defined by the American Diabetes Association criteria. Participants were randomized in a 2:1:1 ratio to 14 weeks of the GLP1R agonist liraglutide, hypocaloric diet, or the dipeptidyl peptidase 4 inhibitor sitagliptin. Treatment with drug was double blind and placebo controlled. Measurements were made at baseline, after 2 weeks of treatment prior to significant anticipated weight loss, and after 14 weeks of treatment. The primary outcomes were measures of endothelial function: flow-mediated vasodilation (FMD), plasminogen activator inhibitor-1 (PAI-1), and urine albumin-to-creatinine ratio (UACR).

**Results:** Ninety-three obese pre-diabetic individuals were randomized, and data from 88 individuals who completed at least a single study day are included in the analysis (liraglutide N=44, diet N=22, sitagliptin N=22). Liraglutide and diet reduced weight and insulin resistance, while sitagliptin did not. There was no significant effect of any treatment on endothelial vasodilator function as measured by FMD (Liraglutide: 2 weeks: difference from baseline +0.70%, 95% CI [-0.71, 2.11], P=0.33, 14 weeks: +1.38% [-0.01, 2.78], P=0.05; Sitagliptin: 2 weeks: +1.92% [-0.05, 3.88], P=0.06, 14 weeks: +1.59% [-0.30, 3.48], P=0.10; Diet: 2 weeks: +1.24% [-0.69, 3.18], P=0.21, 14 weeks: +1.20% [-0.97, 3.38], P=0.28). As baseline endothelial function was normal in the overall cohort, post hoc subgroup analyses were conducted in participants stratified by FMD below or above the median. Individuals with baseline FMD below the median, indicative of endothelial dysfunction, had higher BMI, waist circumference and insulin resistance as compared to those with baseline FMD above the median. All three treatments improved FMD at 2 and 14 weeks in individuals with baseline FMD below the median (P<0.05 for all). Liraglutide reduced PAI-1 at 2 and 14 weeks (2 weeks: -2.3 U/mL [-4.1, -0.6], P<0.01; 14 weeks: -3.7 U/mL [-5.5, -2.0], P<0.001), and diet reduced PAI-1 at 14 weeks (-3.8 U/mL [-6.6, -1.0], P<0.01), but sitagliptin did not change PAI-1 levels. GLP1R antagonism with exendin (9-39) increased fasting blood glucose but did not change FMD or PAI-1 in any treatment group. There was no effect of treatment on UACR. Finally, liraglutide, but not sitagliptin or diet, reduced the chemokine monocyte chemoattractant protein-1 (MCP-1) at 2 and 14 weeks (2 weeks: -10.7 pg/mL [-17.6, -3.7], P<0.01; 14 weeks: -10.7 pg/mL [-17.7, -3.7], P<0.01).

**Conclusions:** Liraglutide and diet cause weight loss, improve insulin resistance and reduce PAI-1 concentrations. Liraglutide, sitagliptin and diet do not change FMD in obese pre-diabetic individuals with normal endothelial function after 14 weeks of treatment. Liraglutide alone lowers the pro-inflammatory and pro-atherosclerotic chemokine MCP-1 at 2 and 14 weeks, indicating that this beneficial cardiovascular effect is independent of weight loss.

**Clinicaltrials.gov Identifier:** NCT03101930

**Research in context:** *What is already known about this subject?:*

- GLP-1 receptor agonists reduce cardiovascular events in patients with type 2 diabetes through unknown mechanism(s).
- GLP-1 receptor agonists cause weight loss in obese individuals with and without diabetes.

*What is the key question?:*

- Are vascular effects of GLP-1 receptor agonists mediated by weight loss or GLP-1 receptor activation?

*What are the new findings?:*

- The GLP-1 receptor agonist liraglutide and diet-induced weight loss reduce circulating PAI-1, a measure of fibrinolytic balance and inflammation, while the DPP-4 inhibitor sitagliptin does not.
- Liraglutide reduces the circulating pro-inflammatory chemokine MCP-1, while sitagliptin and diet-induced weight loss do not.
- Liraglutide, sitagliptin and diet-induced weight loss do not improve flow-mediated dilation, a measure of endothelial vasodilator function, in obese pre-diabetic individuals after 14 weeks of treatment.

*How might this impact on clinical practice in the foreseeable future?:*

- Understanding the mechanisms by which GLP-1 receptor agonists exert vascular effects is critical in improving patient selection and future drug development.

## Introduction

Glucagon-like peptide-1 receptor (GLP1R) agonists decrease cardiovascular morbidity and mortality in patients with type 2 diabetes mellitus (T2DM) [1-3]. GLP1R agonists also cause significant weight loss and have been approved as pharmacologic weight loss therapy [4]. Given that weight loss improves many of the risk factors for cardiovascular disease in patients with T2DM and insulin resistance [5, 6], the question remains whether the beneficial cardiovascular effects of the GLP1R agonists are fully or partially attributable to weight loss.

Dipeptidyl peptidase 4 (DPP4) inhibitors increase endogenous GLP-1 without inducing weight loss. DPP4 inhibitors have not been shown to reduce cardiovascular mortality in T2DM [7]. In addition to preventing the degradation of GLP-1, DPP4 inhibitors also prevent the degradation and formation of a variety of vasoactive peptides, rendering their actions less specific [8].

To understand the weight loss versus GLP1R-dependent effects of the GLP1R agonist liraglutide on vascular function, we compared the effects of liraglutide to hypocaloric diet-induced weight loss and to treatment with the DPP4 inhibitor sitagliptin in a randomized controlled trial. We enrolled obese pre-diabetic individuals and measured weight, metabolic parameters, hemodynamic variables, endothelial function, fibrinolysis, and markers of inflammation at baseline, after two weeks of therapy before anticipated clinically significant weight loss, and after 14 weeks of therapy. We evaluated endothelial function using flow-mediated vasodilation (FMD) and assessed fibrinolysis by measuring plasminogen activator inhibitor-1 (PAI-1), the major inhibitor of tissue-type plasminogen activator *in vivo*. We also evaluated a marker of vascular inflammation by measuring the chemokine monocyte chemoattractant protein-1 (MCP-1). In addition, to assess whether the effects of treatment were GLP1R-dependent, we studied a subset of participants after treatment with the GLP1R antagonist exendin (9-39) and matching vehicle in a randomized, crossover design.

## Methods

### Participants

Men and women aged 18 to 65 years with obesity (BMI ≥ 30 kg/m^2^) and pre-diabetes were eligible. Pre-diabetes was defined using the American Diabetes Association criteria as either impaired fasting glucose between 100-125 mg/dL or impaired glucose tolerance after a 75-gram glucose challenge of 140-199 mg/dL or hemoglobin A1c (HbA1C) 5.7-6.4%. Pregnancy was excluded in women of child-bearing potential by urine β-human chorionic gonadotropin. Individuals with type 1 or type 2 diabetes, resistant hypertension, history of pancreatitis, significant cardiovascular disease, asthma with regular inhaler use, and impaired kidney or liver function were excluded. The study was approved by the Vanderbilt Institutional Review Board, registered at clinicaltrials.gov NCT03101930, and conducted according to the Declaration of Helsinki. All participants provided written informed consent.

### Protocol

Volunteers reported to the Vanderbilt Clinical Research Center to undergo screening. We obtained a medical history, completed a physical examination, collected blood for HbA1C, complete blood count, basic metabolic panel, hepatic function panel, lipid profile, and collected urine for spot urine albumin-to-creatinine ratio (UACR). After an overnight fast, we performed a 75-gram oral glucose tolerance test with samples drawn at 0, 30, 60, 90 and 120 minutes. We measured resting energy expenditure using indirect calorimetry with a ventilated canopy device (CPX/D system; Medical Graphics Corporation, St. Paul, MN) [9] and will evaluate those data in a separate manuscript.

Participants underwent a six-week run-in phase to optimize their medical management (**Supplementary Figure S1A** and detailed methods in **Electronic Supplementary Materials**). After the run-in phase, participants underwent a baseline study day during which we made anthropometric and hemodynamic measurements, measured FMD, collected blood for measurement of glucose, insulin, PAI-1, MCP-1, P-selectin (a measure of platelet and endothelial cell activation), and collected urine for albumin and creatinine.

Participants were then randomized in parallel to liraglutide 1.8mg/day (Novo Nordisk), sitagliptin 100mg/day (Merck and Co, Inc), or hypocaloric diet in a 2:1:1 ratio, stratified by race. Within each stratum, we used a block randomization algorithm with a block size of four. A higher proportion of participants were randomized to liraglutide to enable future investigation of individual predictors of response. Liraglutide was given on a dose escalation starting at 0.6mg/day for week 1, 1.2mg/day for week 2 and then up to the full dose of 1.8mg/day at the start of week 3. Treatment with liraglutide or sitagliptin was double blind and placebo-controlled, while treatment with diet was unblinded. Liraglutide and matching placebo were a generous gift from Novo Nordisk.

In the context of the COVID-19 pandemic and after approval from the Data Safety Monitoring Board and the Institutional Review Board, we modified the randomization ratio to enrich for assignment to liraglutide and diet on September 29, 2020.

Participants were treated for a total of 14 weeks. The first 74 participants underwent four study days in a 2×2 crossover study of the GLP1R antagonist exendin (9-39) and placebo. The first two study days were conducted after two weeks of treatment to measure the short-term effects of liraglutide, sitagliptin and diet. The first and second study days were separated by 48 hours and participants received a single dose infusion of placebo or exendin (9-39) on each day to assess the contribution of GLP1R activation to any observed effects. We randomly assigned individuals to two sequences (exendin → placebo or placebo → exendin) in a 1:1 ratio using a block randomization algorithm with a block size of two. The third and fourth study days were completed after 14 weeks to measure the effects of sustained liraglutide, sitagliptin and diet-induced weight loss. The third and fourth study days were also separated by 48 hours and participants again received placebo or exendin (9-39) infusion in random order. The last 14 individuals studied did not participate in the crossover study with exendin (9-39) due to lack of drug availability, and only underwent two study days, one after 2 weeks and one after 14 weeks of treatment. Placebo vehicle was infused during each of these study days. All participants underwent repeat urine collection after 13.5 weeks at a separate visit to the Clinical Research Center.

The study biostatistician prepared the R programs for generating the allocation schedules. A biostatistician unrelated to the study and the investigators ran the programs using a different random number generator seed to generate the allocation schedule and maintain the investigators’ blinded status. Vanderbilt Investigational Drug Services maintained the allocation schedules, and stored, prepared, and labeled all medications.

### Study Day Procedures

Participants randomized to drug were called the night prior to each study day to ensure they had taken their subcutaneous injection. On the morning of each study day, participants arrived at the Clinical Research Center after an overnight fast. Those randomized to drug took their oral medication upon arrival (**Supplementary Figure S1B**). Participants rested supine for 45 minutes, and we measured blood pressure and heart rate and collected blood for PAI-1, MCP-1, and P-selectin. After this rest period, participants were given placebo or the GLP1R antagonist exendin-(9-39) (Clinalfa®, Bachem Distribution Services; Weil am Rhein, Germany; IND #122,217) in randomized order on study days 1 and 2, and on study days 3 and 4. Exendin-(9-39) was given as an IV bolus of 7500 pmol/kg over one minute followed by continuous infusion of 350 pmol/kg/min for the remainder of the study. Infusion with placebo or exendin-(9-39) was double blind.

An hour after the start of the infusion, we measured FMD, collected blood for measurement of PAI-1, fasting glucose, and insulin. We then administered sublingual nitroglycerin 0.4mg for measurement of endothelium-independent nitroglycerin-mediated vasodilation (NMD).

A mixed meal study was performed after vascular studies were completed and the results of the mixed meal study will be presented in a separate manuscript.

### Measurement of Flow-Mediated Vasodilation

Measurement of endothelium-dependent and endothelium-independent vasodilation was performed in a quiet, temperature-controlled (23°C) room. Participants were asked to refrain from alcohol and caffeine for at least 12 hours prior to vascular function measurements. In addition, participants were asked to abstain from phosphodiesterase type 5 inhibitors for at least one week as these drugs may confound the measurement of vascular function or cause precipitous hypotension after nitroglycerin. FMD and NMD were measured as reported previously and detailed in **Electronic Supplementary Materials**.

### Hypocaloric Diet

Participants in the hypocaloric diet arm were given a caloric goal designed to achieve a weight loss similar to that expected in the liraglutide treatment arm; based on prior studies of 1.8 mg/d liraglutide this was predicted to be ∼0.27 kg/week [10]. Each participant’s caloric goal was determined by measuring resting energy expenditure at baseline and calculated to reduce total energy balance by 390 kcal/d [11]. Participants were provided counseling and written instructions on how to achieve their daily caloric goal, including use of their own mobile phone applications to monitor caloric intake. To assure compliance with the prescribed caloric goal, participants met with the study dietitian every other week for problem solving and review of diet intake logs.

### Laboratory Analyses

A YSI glucose analyzer (YSI Life Sciences, Yellow Springs, OH) was used to measure plasma glucose immediately after collection. All remaining samples were stored at -80°C in aliquots until the time of assay. Plasma insulin samples were collected in tubes containing aprotinin, and insulin was measured by radioimmunoassay (EMD Millipore, Billerica, MA). The assay cross-reacts with 38% intact proinsulin, but not with C-peptide (≤ 0.01%). Blood for PAI-1 was collected in 0.105 M acidified sodium citrate and samples were analyzed using commercially available two-site ELISA using chromogenic substrates (TriniLIZE, Trinity Biotech, Berkeley Heights, NJ and American Diagnostica Inc, Stamford, CT). Plasma samples for MCP-1 were collected in a BD potassium EDTA purple top vacutainer tube, spun immediately at 3,000 RPM for 10 minutes, then aliquoted for storage at -80C until measured by immunoassay (Meso Scale Diagnostics, Rockville, MD). Serum samples for p-selectin were collected in a BD red top vacutainer tube, incubated at room temperature for 30 minutes, spun at 3,000 RPM for 10 minutes, then aliquoted for storage at -80C until measured by ELISA (R&D Systems, Minneapolis, MN). Creatinine was measured using the Jaffe method. Urine albumin was measured by turbidimetric immunoassay with endpoint determination. HbA1C was measured using high pressure liquid chromatography.

### Statistical Analyses

The primary endpoints were endothelial vascular function (FMD, UACR) and fibrinolytic function (PAI-1) after 2 weeks (prior to clinically significant weight loss) and after 14 weeks of treatment. Secondary endpoints reported here include blood pressure, heart rate, fasting insulin and glucose, MCP-1, and P-selectin. Endpoints related to the mixed meal study will be analyzed and reported separately. All analyses were by original assigned groups.

Descriptive statistics of patient baseline characteristics are presented as mean ± SD for continuous variables and frequencies and proportions for categorical variables. Between-group comparison was performed using Kruskal–Wallis test or Pearson’s chi-squared test. To evaluate the treatment effects on weight, fasting blood glucose, systolic blood pressure, diastolic blood pressure, heart rate and pre-infusion PAI-1, P-selectin, and MCP-1, separate multivariable generalized least squares linear regression models were fitted using the data from vehicle-only study days. Treatment (liraglutide, sitagliptin or diet), time (2- or 14-week), baseline measurement as well as interaction between treatment and time were included as independent variables. A compound symmetry structure for within-subject correlation was used. For fasting blood glucose, fasting insulin, Homeostatic Model Assessment for Insulin Resistance (HOMA-IR), forearm blood flow measures including FMD percent, and post-infusion PAI-1, generalized least squares linear regression models were fitted using data from all study days. Treatment (liraglutide, sitagliptin or diet), infusion (vehicle or exendin (9-39)), time (2- or 14-week), baseline measurement as well as all the two-way and three-way interactions between treatment, time and infusion were included as independent variables. For all forearm blood flow measures, gender was included as a covariate. Inferences on the contrasts of interest were conducted using Wald test. Estimates of change in outcome from baseline to 2- and 14-weeks were calculated based on the multivariable regression model. Spearman’s rank correlation was computed to assess the relationship between PAI-1 and HOMA-IR or MCP-1. All the analyses were performed using the statistical software R 4.1.0.

We powered our study based on anticipated improvement in endothelial function. The original sample size was planned at 160 individuals to allow for stratified analyses by race, gender and GLP1R rs6923761 single nucleotide polymorphism genotype. Due to recruitment shortfalls resulting from the COVID-19 pandemic, and in consultation with our Data Safety Monitoring Board and our study biostatistician, we stopped the study after completion of 88 individuals based on the following post hoc power calculation. In studies of weight loss, improvement in endothelial function is proportionate to weight loss and the degree of weight loss achieved with liraglutide in the LEADER study has been associated with a 1.3%±2.1% increase in brachial artery FMD [12]. To detect a difference of 1.3%±2.1% for within-individual comparisons, we had 79% power in the diet and sitagliptin arms (N=22), and 98% power in the liraglutide arm (N=44).

## Results

### Participant characteristics

Participant recruitment and follow-up was completed between May 2017 to June 2021. Ninety-three individuals were randomized to treatment, with 46 to liraglutide, 23 to sitagliptin, and 24 to hypocaloric diet (**Supplementary Figure S2**). Three participants dropped out prior to receiving treatment, and two dropped out after receiving treatment but before completing any study days. Data from the remaining 88 individuals (44 randomized to liraglutide, 22 to sitagliptin, and 22 to hypocaloric diet) were analyzed. Seven individuals dropped out after completing study days 1 and 2 (two randomized to liraglutide, five to diet) and their available data were included in the analyses. As shown in **Table 1**, baseline participant characteristics were similar across treatment arms.

**Table 1.** Baseline Participant Characteristics.

| Measures | Liraglutide<br>N=44 | Sitagliptin<br>N=22 | Diet<br>N=22 | Combined<br>N=88 |
| --- | --- | --- | --- | --- |
| Age, years | 49.8±10.1 | 52.4±10.5 | 49.2±12.5 | 50.3±10.8 |
| Gender |  |  |  |  |
| Male | 29.5% (13) | 31.8% (7) | 36.4% (8) | 31.8% (28) |
| Female | 70.5% (31) | 68.2% (15) | 63.6% (14) | 68.2% (60) |
| Race |  |  |  |  |
| Asian | 4.5% (2) | 0% (0) | 0% (0) | 2.3% (2) |
| Black or African American | 9.1% (4) | 13.6% (3) | 18.2% (4) | 12.5% (11) |
| White | 86.4% (38) | 81.8% (18) | 77.3% (17) | 83.0% (73) |
| More Than One Race | 0% (0) | 4.5% (1) | 4.5% (1) | 2.3% (2) |
| Ethnicity |  |  |  |  |
| Hispanic or Latino | 2.3% (1) | 9.1% (2) | 4.5% (1) | 4.5% (4) |
| NOT Hispanic or Latino | 95.5% (42) | 90.9% (20) | 95.5% (21) | 94.3% (83) |
| Unknown / Not Reported | 2.3% (1) | 0% (0) | 0% (0) | 1.1% (1) |
| Weight, kg | 108.8±20.9 | 111.4±22.0 | 111.3±21.5 | 110.1±21.1 |
| BMI, kg/m<sup>2</sup> | 38.8±6.1 | 39.9±6.0 | 38.4±5.9 | 39.0±6.0 |
| Waist Circumference, cm | 115.8±11.5 | 118.4±14.5 | 117.5±13.6 | 116.9±12.8 |
| Hip Circumference, cm | 126.2±12.7 | 126.2±12.0 | 125.7±10.9 | 126.1±11.9 |
| Fasting blood glucose, mg/dL | 96.2±10.1 | 100.2±9.4 | 96.7±11.8 | 97.4±10.4 |
| Fasting insulin, µU/mL | 21.4±13.9 | 26.9±14.0 | 18.4±7.4 | 22.3±12.8 |
| OGTT 2-hour blood glucose, mg/dL | 142.0±24.8 | 148.1±41.1 | 130.2±27.8 | 140.6±30.7 |
| HOMA-IR | 5.0±3.0 | 6.5±3.3 | 4.6±2.4 | 5.3±3.0 |
| HOMA2 | 0.41±0.25 | 0.52±0.27 | 0.36±0.15 | 0.43±0.24 |
| Hemoglobin A1c, % | 5.7±0.3 | 5.8±0.3 | 5.7±0.3 | 5.7±0.3 |
| Total cholesterol, mg/dL | 191.0±39.2 | 192.5±31.2 | 170.9±49.9 | 186.6±40.7 |
| Triglycerides, mg/dL | 122.2±51.4 | 143.3±59.6 | 115.0±67.1 | 125.9±58.0 |
| HDL, mg/dL | 47.5±9.6 | 45.6±11.2 | 44.1±10.9 | 46.2±10.3 |
| LDL, mg/dL | 119.1±31.9 | 118.2±27.0 | 111.5±35.7 | 117.0±31.4 |
| Systolic blood pressure, mmHg | 124.1±7.7 | **120.2±11.3\*** | 127.7±8.3 | 124.1±9.1 |
| Diastolic blood pressure, mmHg | 77.6±9.3 | 74.7±8.7 | 77.8±7.0 | 76.9±8.6 |
| Heart rate, bpm | 64.9±7.4 | 67.2±9.0 | 63.8±8.8 | 65.2±8.2 |
| Anti-hypertensive agent use | 25.0% (11) | 36.4% (8) | 45.5% (10) | 33.0% (29) |
| FMD, % | 10.54±5.21 | 10.39±5.37 | 10.22±5.25 | 10.42±5.20 |

All measures shown as mean±SD for continuous variables and % (N) for categorical variables.
OGTT indicates oral glucose tolerance test; HOMA-IR, Homeostatic Model Assessment for Insulin Resistance; HOMA2, Homeostatic Model Assessment 2. Asterisk (\*) indicates P<0.05 versus diet using Wilcoxon rank-sum test.

### Effect of treatment on weight and fasting metabolic measures

Liraglutide and diet caused weight loss, while sitagliptin did not (**Figure 1A** and **Table 2**). Individuals in the diet arm lost significant weight from baseline at both 2 and 14 weeks (2 weeks: difference -1.4 kg, 95% CI [-2.4, -0.3], P=0.01; 14 weeks: -4.9 kg [-6.1, -3.7], P<0.001). Individuals in the liraglutide arm lost a small amount of weight from baseline to 2 weeks (−0.5 kg [-1.3, 0.3], P=0.20), and continued to lose significant weight by 14 weeks (−2.7 kg [-3.5, -1.9], P<0.001). Hypocaloric diet-treated participants lost more weight compared to liraglutide-treated participants at 14 weeks (−2.3 kg [-3.7, -0.9], P<0.01). In addition, both diet-treated and liraglutide-treated participants lost more weight than sitagliptin-treated participants at 14 weeks (Liraglutide vs Sitagliptin -2.0 kg [-3.3, -0.6], P<0.01; Diet vs Sitagliptin -4.2 kg [-5.8, -2.6], P<0.001).

**Figure 1.**
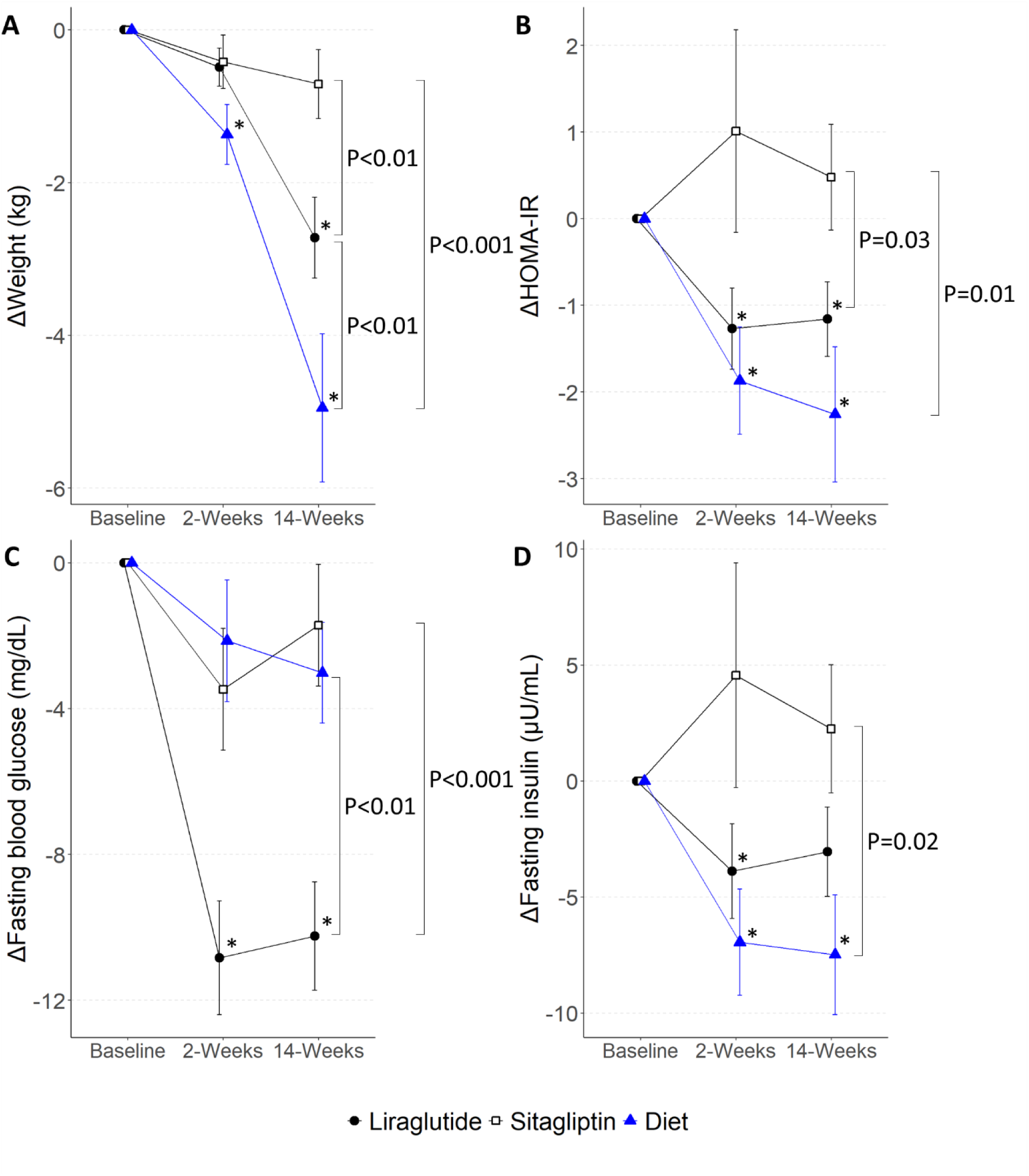
The Effect of Treatment on Weight, Fasting Blood Glucose, Fasting Insulin, and Insulin Resistance. Plots show mean ± SEM for **(A)** weight, **(B)** HOMA-IR, **(C)** fasting blood glucose, and **(D)** fasting insulin at 2 and 14 weeks of treatment as change from baseline. Asterisk (*) symbols indicate P<0.05 for estimates of change from baseline, and brackets indicate difference between treatments at 14 weeks. HOMA-IR indicates Homeostatic Model Assessment of Insulin Resistance.

**Table 2.** Effects of Treatment.

| Measures | <u>Liraglutide</u> |  |  | <u>Sitagliptin</u> |  |  | <u>Diet</u> |  |  |
| --- | --- | --- | --- | --- | --- | --- | --- | --- | --- |
|  | Baseline | 2 weeks | 14 weeks | Baseline | 2 weeks | 14 weeks | Baseline | 2 weeks | 14 weeks |
| Weight, kg | 108.8±20.9 | 108.3±21.0 | **106.4±22.1\***†‡ | 111.4±22.0 | 111.0±22.6 | **110.7±22.3‡** | 111.3±21.5 | **109.9±21.4\*** | **108.6±18.1\*** |
| Fasting glucose, mg/dL | 95.3±8.6 | **84.3±7.9\***†‡ | **85.2±7.3\***†‡ | 97.6±10.0 | 93.9±8.1 | 96.6±5.6 | 94.5±12.0 | 92.4±11.3 | 91.2±9.8 |
| Fasting insulin, µU/mL | 22.7±16.8 | **18.3±12.5\***† | 20.3±14.7 | 23.3±14.4 | **29.4±25.4‡** | **26.0±19.0‡** | 26.7±21.2 | **19.7±16.5\*** | **20.3±13.7\*** |
| HOMA-IR | 5.4±4.0 | **3.9±2.8\***† | **4.4±3.4\***† | 5.5±3.3 | **6.9±6.0‡** | **6.1±4.3‡** | 6.6±6.1 | **4.8±4.6\*** | **4.8±3.7\*** |
| Systolic BP, mmHg | 124.1±7.7 | 122.9±6.3 | **122.2±7.8‡** | 120.2±11.3 | 117.5±11.3 | 118.2±13.9 | 127.7±8.3 | **121.7±6.8\*** | **119.7±11.1\*** |
| Diastolic BP, mmHg | 77.6±9.3 | **78.3±7.7†** | **77.7±7.1†‡** | 74.7±8.7 | 72.3±7.8 | 73.1±9.8 | 77.9±7.0 | 76.5±5.1 | **74.8±8.3\*** |
| Heart rate, bpm | 64.9±7.5 | **69.0±6.4\***†‡ | **68.9±5.6\***†‡ | 67.2±9.0 | 66.2±9.2 | 65.9±8.5 | 63.8±8.8 | 63.2±9.5 | **61.7±7.9\*** |
| PAI-1, U/mL | 20.2±9.2 | **17.0±6.4\*** | **16.9±6.4\***† | 18.5±8.6 | 16.9±6.5 | **19.5±7.0‡** | 18.5±8.0 | 19.4±7.5 | **15.3±5.9\*** |
| MCP-1, pg/mL | 105.4±36.1 | **95.0±25.0\*** | **95.2±26.9\*** | 107.5±25.8 | 107.6±30.8 | 106.7±26.0 | 108.2±34.1 | 103.1±36.9 | 109.2±34.3 |
| P-selectin, ng/mL | 57.1±23.0 | 56.6±24.1 | 52.9±20.9 | 52.3±25.3 | 51.6±22.3 | 51.8±19.5 | 58.6±22.0 | 57.5±28.4 | 60.8±22.1 |
| UACR | 12.0±23.6 | ND | 10.5±14.8 | 7.9±7.6 | ND | 9.2±10.7 | 6.3±3.8 | ND | 10.1±19.4 |
| **Forearm Blood Flow Measures** |  |  |  |  |  |  |  |  |  |
| Baseline diameter, mm | 3.35±0.51 | 3.25±0.47 | 3.33±0.55 | 3.51±0.62 | 3.47±0.63 | 3.47±0.66 | 3.41±0.67 | 3.44±0.74 | 3.40±0.71 |
| FMD, % | 10.54±5.21 | 11.70±5.14 | 12.01±6.18 | 10.39±5.37 | 12.18±4.56 | 11.98±4.45 | 10.22±5.25 | 10.99±5.12 | 10.73±4.80 |
| Pre-nitro diameter, mm | 3.39±0.51 | 3.14±0.40 | 3.20±0.42 | 3.82±0.65 | 3.38±0.80 | 3.68±0.72 | 3.46±0.77 | 3.55±0.85 | 3.88±0.86 |
| NMD, % | 20.39±8.59 | 22.32±8.61 | 21.60±10.07 | 19.31±6.39 | 26.57±14.19 | 21.92±8.23 | 21.05±9.92 | 20.09±9.27 | 15.44±10.91 |

All measures shown as mean±SD. HOMA-IR indicates Homeostatic Model Assessment for Insulin Resistance; PAI-1, plasminogen activator inhibitor-1; MCP-1, monocyte chemoattractant protein-1; UACR, urine albumin-to-creatinine ratio; FMD, flow-mediated dilation; Nitro, nitroglycerin; NMD, nitroglycerin-mediated dilation. ND indicates not done. Asterisk (\*) indicates P<0.05 versus baseline; dagger (†) indicates P<0.05 versus sitagliptin; double dagger (‡) P<0.05 versus diet.

Liraglutide and diet decreased HOMA-IR, a measure of insulin resistance, while sitagliptin did not (**Figure 1B** and **Table 2**). Diet caused the greatest decrease in HOMA-IR at both 2 and 14 weeks compared to baseline (2 weeks: -1.9 [-3.1, -0.7], P<0.01; 14 weeks: -2.4 [-3.7, -1.1], P<0.001). Liraglutide also decreased HOMA-IR at both 2 and 14 weeks (2 weeks: -1.3 [-2.1, -0.4], P<0.01; 14 weeks: -1.2 [-2.0, -0.3], P<0.01). Both diet-treated and liraglutide-treated participants had a greater decrease in HOMA-IR than sitagliptin-treated participants at 14 weeks (Liraglutide vs Sitagliptin -1.6 [-3.1, -0.1], P = 0.03; Diet vs Sitagliptin -2.3 [-4.1, -0.5], P=0.01).

As shown in **Figure 1C** and **Table 2**, liraglutide decreased fasting blood glucose, while sitagliptin and hypocaloric diet did not. Liraglutide caused an early and sustained decrease in fasting blood glucose from baseline (2 weeks: -10.9 mg/dL [-13.5, -8.2], P<0.001; 14 weeks: - 10.3 mg/dL [-13.0, -7.6], P<0.001). Sitagliptin and hypocaloric diet did not significantly alter fasting blood glucose at 2 or 14 weeks (Sitagliptin: 2 weeks: -3.3 mg/dL [-7.1, 0.5], P=0.09; 14 weeks: -2.1 mg/dL [-5.9, 1.7], P=0.27; Diet: 2 weeks: -2.1 mg/dL [-5.9, 1.6], P=0.26; 14 weeks: - 2.9 mg/dL [-7.0, 1.3], P=0.18). Liraglutide caused the greatest reduction in fasting glucose at 14 weeks (Liraglutide vs Sitagliptin -9.2 mg/dL [-13.9, -4.5], P<0.001; Liraglutide vs Diet -6.9 mg/dL [-11.9, -2.0], P<0.01).

Finally, as shown in **Figure 1D** and **Table 2**, diet decreased fasting insulin at both 2 and 14 weeks compared to baseline (2 weeks: -7.0 µU/mL [-11.6, -2.3], P<0.01; 14 weeks: -8.2 µU/mL [-13.3, -3.1], P<0.01). Liraglutide significantly decreased fasting insulin at 2 but not 14 weeks (2 weeks: -3.7 µU/mL [-7.1, -0.4], P=0.03; 14 weeks: -3.1 µU/mL [-6.4, 0.3], P=0.08). Sitagliptin did not alter fasting insulin at 2 or 14 weeks.

### Effect of treatment on endothelial vasodilator function

Baseline brachial artery diameter and FMD percent were comparable among treatment groups (**Figure 2A** and **Table 2**). Liraglutide, sitagliptin and hypocaloric diet did not significantly change FMD at 2 or 14 weeks compared to baseline (Liraglutide: 2 weeks: +0.70% [-0.71, 2.11], P=0.33, 14 weeks: +1.38% [-0.01, 2.78], P=0.05; Sitagliptin: 2 weeks: +1.92% [-0.05, 3.88], P=0.06, 14 weeks: +1.59% [-0.30, 3.48], P=0.10; Diet: 2 weeks: +1.24% [-0.69, 3.18], P=0.21, 14 weeks: +1.20% [-0.97, 3.38], P=0.28). There was no significant change in endothelium-independent NMD in any of the treatment groups (**Table 2**).

**Figure 2.**
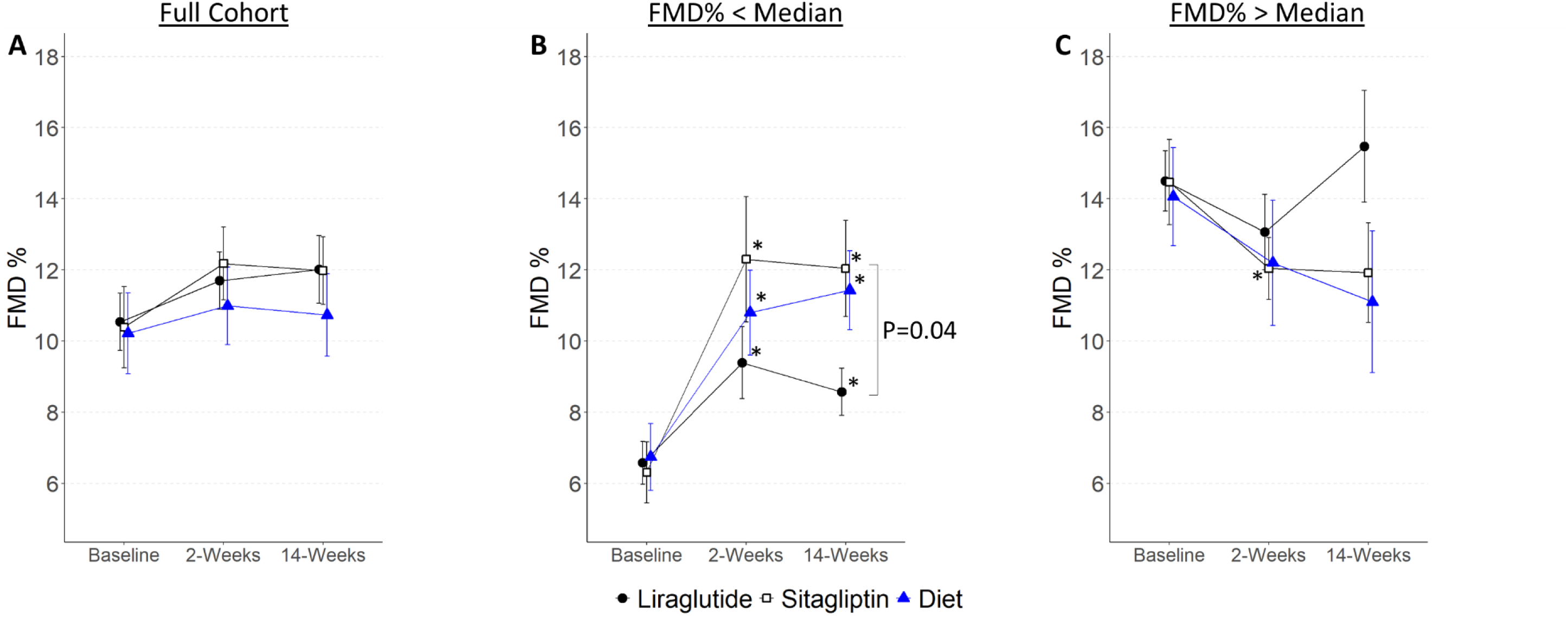
The Effect of Treatment on FMD. Plots show mean ± SEM for **(A)** FMD percent in the entire cohort, **(B)** in subgroup with baseline FMD percent below median for gender, and **(C)** in subgroup with baseline FMD above median for gender. Asterisk (*) symbols indicate P<0.05 for estimates of change from baseline, and brackets indicate difference between treatments at 14 weeks. FMD indicates flow-mediated dilation.

In considering the cohort’s clinical characteristics, we noted that baseline endothelial function was similar to that measured in our prior studies in healthy subjects [13, 14]. In a post hoc exploratory analysis, we divided the cohort into those with baseline FMD below or above the median for gender, to query the effect of treatment in individuals with normal and reduced endothelial function. Baseline characteristics of these subgroups are shown in **Table 3**. Three individuals were missing baseline FMD and were excluded from this analysis. Individuals with lower baseline FMD, indicative of endothelial dysfunction, had increased BMI, waist circumference, fasting insulin and insulin resistance as measured by HOMA-IR and HOMA2 compared to those with higher baseline FMD. As shown in **Figure 2B**, in individuals with lower baseline FMD, treatment with liraglutide, sitagliptin and hypocaloric diet improved FMD after 2 and 14 weeks. Of note, FMD in individuals with normal baseline FMD did not change with any intervention, with the exception of sitagliptin treatment at 2 weeks (**Figure 2C**).

**Table 3.** Participant Characteristics by FMD Subgroups.

| Measures | FMD% | FMD% | Combined |
| --- | --- | --- | --- |
|  | < Median | > Median |  |
|  | N=43 | N=42 |  |
| Age, years | 48.3±10.9 | 51.8±10.7 | 50.0±10.9 |
| Gender |  |  |  |
| Male | 32.6% (14) | 31.0% (13) | 31.8% (27) |
| Female | 67.4% (29) | 69.0% (29) | 68.2% (58) |
| Race |  |  |  |
| Asian | 4.7% (2) | 0% (0) | 2.4% (2) |
| Black or African American | 7.0% (3) | 19.0% (8) | 12.9% (11) |
| White | 88.4% (38) | 76.2% (32) | 82.4% (70) |
| More Than One Race | 0% (0) | 4.8% (2) | 2.4% (2) |
| Ethnicity |  |  |  |
| Hispanic or Latino | 4.7% (2) | 4.8% (2) | 4.7% (4) |
| NOT Hispanic or Latino | 93.0% (40) | 95.2% (40) | 94.1% (80) |
| Unknown / Not Reported | 2.3% (1) | 0% (0) | 1.2% (1) |
| Weight, kg | 113.9±22.7 | 106.0±19.5 | 110.0±21.4 |
| BMI, kg/m<sup>2</sup> | **39.9±5.6\*** | 37.8±6.2 | 38.9±6.0 |
| Waist Circumference, cm | **119.9±13.4\*** | 113.7±11.6 | 117.0±12.9 |
| Hip Circumference, cm | 128.4±12.1 | 123.9±11.6 | 126.3±12.0 |
| Fasting blood glucose, mg/dL | 97.9±9.4 | 96.7±11.6 | 97.3±10.5 |
| Fasting insulin, μU/mL | **27.0±11.9\*** | 18.7±12.8 | 22.2±13.0 |
| OGTT 2-hour blood glucose, mg/dL | 146.8±32.3 | 136.3±29.4 | 141.3±31.0 |
| HOMA-IR | **6.7±3.0\*** | 4.3±2.7 | 5.3±3.0 |
| HOMA2 | **0.52±0.23\*** | 0.35±0.23 | 0.42±0.24 |
| Hemoglobin A1c, % | 5.7±0.3 | 5.7±0.3 | 5.7±0.3 |
| Total cholesterol, mg/dL | 185.4±30.1 | 190.3±44.3 | 187.9±37.8 |
| Triglycerides, mg/dL | 120.4±54.2 | 133.0±62.2 | 126.7±58.3 |
| HDL, mg/dL | 45.8±10.7 | 46.0±9.9 | 45.9±10.3 |
| LDL, mg/dL | 114.9±26.8 | 117.8±36.2 | 116.4±31.8 |
| Systolic blood pressure, mmHg | 123.4±8.1 | 124.8±9.8 | 124.1±9.0 |
| Diastolic blood pressure, mmHg | 76.0±8.9 | 77.8±8.7 | 76.9±8.8 |
| Heart rate, bpm | 64.7±8.7 | 65.6±8.0 | 65.2±8.3 |
| PAI-1, U/mL | 20.9±8.8 | 17.6±8.5 | 19.3±8.8 |
| MCP-1, pg/mL | 106.5±34.8 | 109.1±31.4 | 107.8±33.0 |
| FMD, % | **6.55±2.81\*** | 14.39±3.94 | 10.42±5.20 |

All measures shown as mean ± SD for continuous variables and % (N) for categorical variables. OGTT indicates oral glucose tolerance test; HOMA-IR, Homeostatic Model Assessment for Insulin Resistance; HOMA2, Homeostatic Model Assessment 2; PAI-1, plasminogen activator inhibitor-1; MCP-1, monocyte chemoattractant protein-1; FMD, flow-mediated dilation. \* P<0.05 versus FMD% greater than median using Wilcoxon rank-sum test.

Urine albumin-to-creatinine ratio, which is associated with vascular endothelial function and is a predictor of cardiovascular events [15-17], was not changed by treatment (**Table 2**).

### Effect of treatment on circulating measures of fibrinolysis and inflammation

We measured concentrations of PAI-1, the major inhibitor of endogenous fibrinolysis and a marker of inflammation. Liraglutide significantly decreased PAI-1 concentration from baseline to 2 and 14 weeks (2 weeks: -2.3 U/mL [-4.1, -0.6], P<0.01; 14 weeks: -3.7 U/mL [-5.5, -2.0], P<0.001; **Figure 3A**). Hypocaloric diet significantly decreased PAI-1 concentration from baseline to 14 weeks (−3.8 U/mL [-6.6, -1.0], P<0.01). Sitagliptin treatment did not affect PAI-1 concentration. In addition, PAI-1 levels were lower at 14 weeks in both the liraglutide- and diet-treated individuals compared to sitagliptin (Liraglutide vs Sitagliptin: -3.7 U/mL [-6.7, -0.6], P=0.02; Diet vs Sitagliptin: -4.8 U/mL [-8.5, -1.1], P=0.01). Finally, PAI-1 concentrations correlated with HOMA-IR both at baseline (r=0.34, P<0.01) and after treatment (Liraglutide r=0.30, P<0.01; Sitagliptin r=0.32, P<0.05; Diet r=0.49, P<0.01).

**Figure 3.**
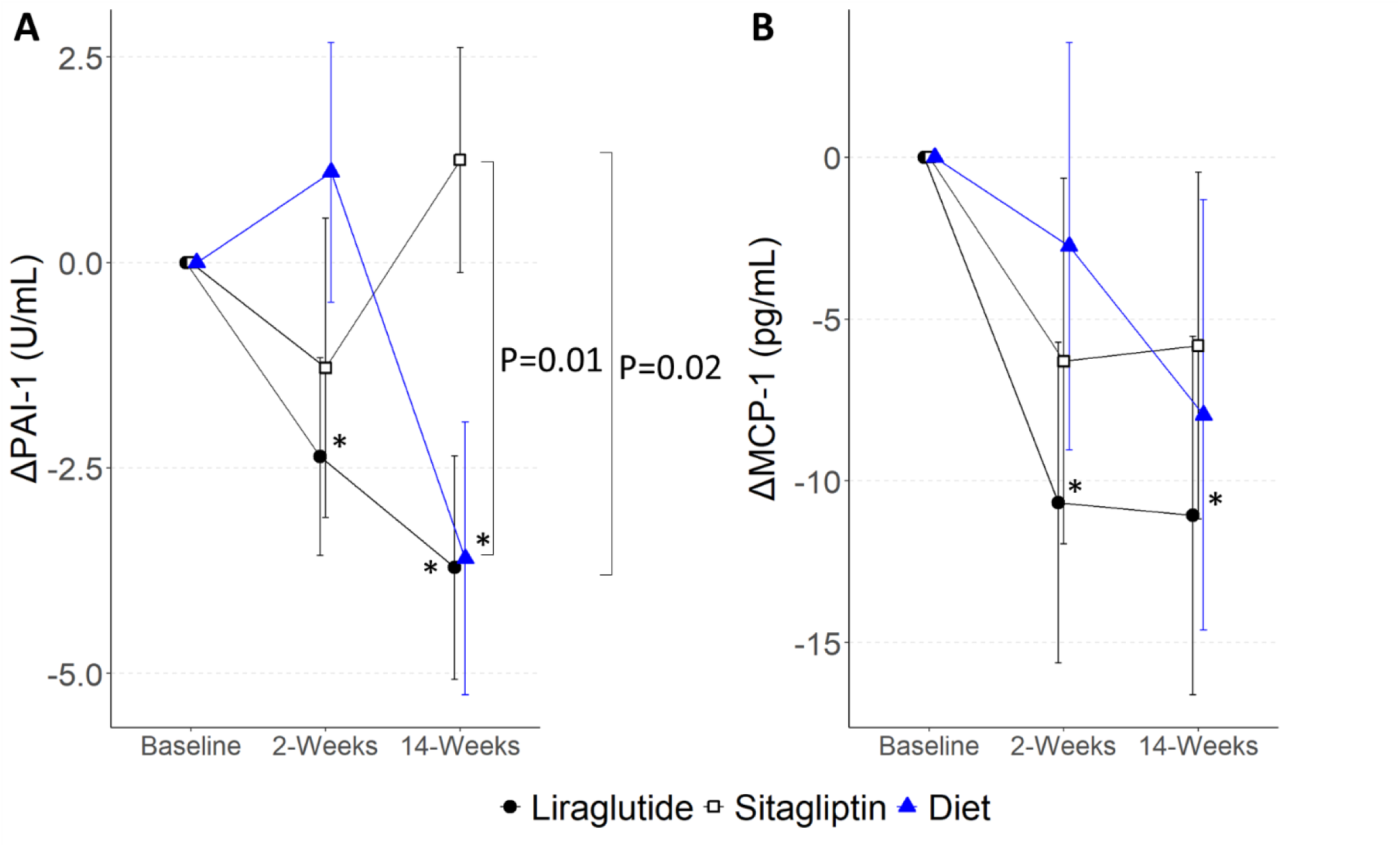
The Effect of Treatment on PAI-1 and MCP-1. Plots show mean ± SEM for **(A)** PAI-1 and **(B)** MCP-1 at 2 and 14 weeks of treatment as change from baseline. Asterisk (*) symbols indicate P<0.05 for estimates of change from baseline, and brackets indicate difference between treatments at 14 weeks. PAI-1 indicates plasminogen activator inhibitor-1; MCP-1, monocyte chemoattractant protein-1.

We next measured concentrations of the circulating chemokine MCP-1. Liraglutide decreased MCP-1 at both 2 and 14 weeks (2 weeks: -10.7 pg/mL [-17.6, -3.7], P<0.01; 14 weeks: -10.7 pg/mL [-17.7, -3.7], P<0.01; **Figure 3B**). Sitagliptin and hypocaloric diet did not significantly change MCP-1 levels. MCP-1 levels did not correlate with HOMA-IR (data not shown) but correlated with PAI-1 at baseline (r=0.30, P=0.01) and after treatment with liraglutide at 2 weeks (r=0.32, P<0.05), but not 14 weeks (r=0.08, P=0.60). Finally, P-selectin, a marker of platelet and endothelial activation, was unchanged by treatment (**Table 2**).

### Effect of treatment on resting hemodynamics

Systolic blood pressure decreased significantly after hypocaloric diet (2 weeks: -6.0 mmHg [-9.4, -2.7], P<0.001; 14 weeks: -8.0 mmHg [-11.6, -4.3], P<0.001), but not with liraglutide or sitagliptin (**Supplementary Figure S3A**). Diastolic blood pressure and heart rate also decreased significantly after 14 weeks of hypocaloric diet (DBP: -3.8 mmHg [-6.7, -0.9], P=0.01; HR: -2.9 bpm [-5.4, -0.4], P=0.02; **Supplementary Figure S3B**). Liraglutide treatment increased heart rate at both 2 weeks (+4.0 bpm [2.4, 5.7], P<0.001) and 14 weeks (+3.7 bpm [2.1, 5.4], P<0.001; **Supplementary Figure S3C**).

### Effect of GLP1R antagonism

To test the hypothesis that GLP1R activation contributes to the effects of liraglutide or sitagliptin, we pre-treated participants with the GLP1R antagonist exendin (9-39) and vehicle at 2 and 14 weeks. As shown in **Figure 4A** and **Table 4**, GLP1R antagonism with exendin (9-39) raised fasting blood glucose in all treatment groups at both 2 and 14 weeks. Overall, exendin did not change FMD in liraglutide, sitagliptin, or diet-treated participants (**Figure 4B**). In the stratified analysis of those with reduced FMD, exendin decreased FMD in sitagliptin-treated individuals at 2 weeks (−3.6 [-7.0, -0.3], P=0.03), but not at 14 weeks (−1.0 [-4.3, 2.3], P=0.56) as compared to vehicle (**Figure 4C**). Finally, there was no acute effect of GLP1R antagonism on PAI-1 concentrations in any treatment group (**Table 4**).

**Figure 4.**
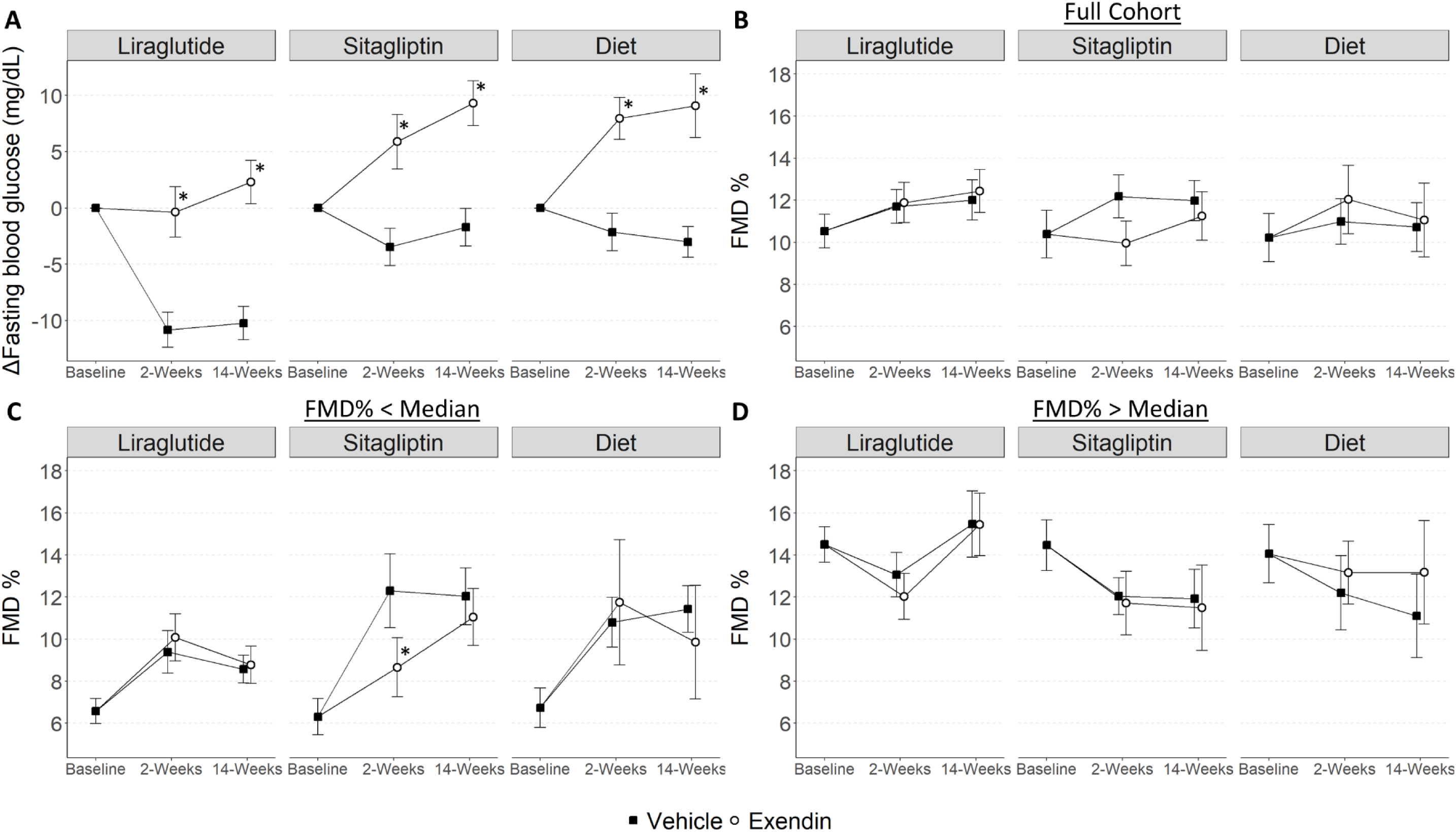
The Effect of the GLP1R Antagonist Exendin (9-39) on Fasting Blood Glucose and FMD. Plots show mean ± SEM for **(A)** fasting blood glucose and **(B-D)** FMD percent. **(A)** Fasting blood glucose at 2 and 14 weeks of treatment as change from baseline. **(B)** FMD percent in the entire cohort; **(C)** in subgroup with baseline FMD percent below median; **(D)** in subgroup with baseline FMD above median. Asterisk (*) symbols indicate P<0.05 exendin versus vehicle. GLP1R indicates glucagon-like peptide-1 receptor; FMD, flow-mediated dilation.

**Table 4.**
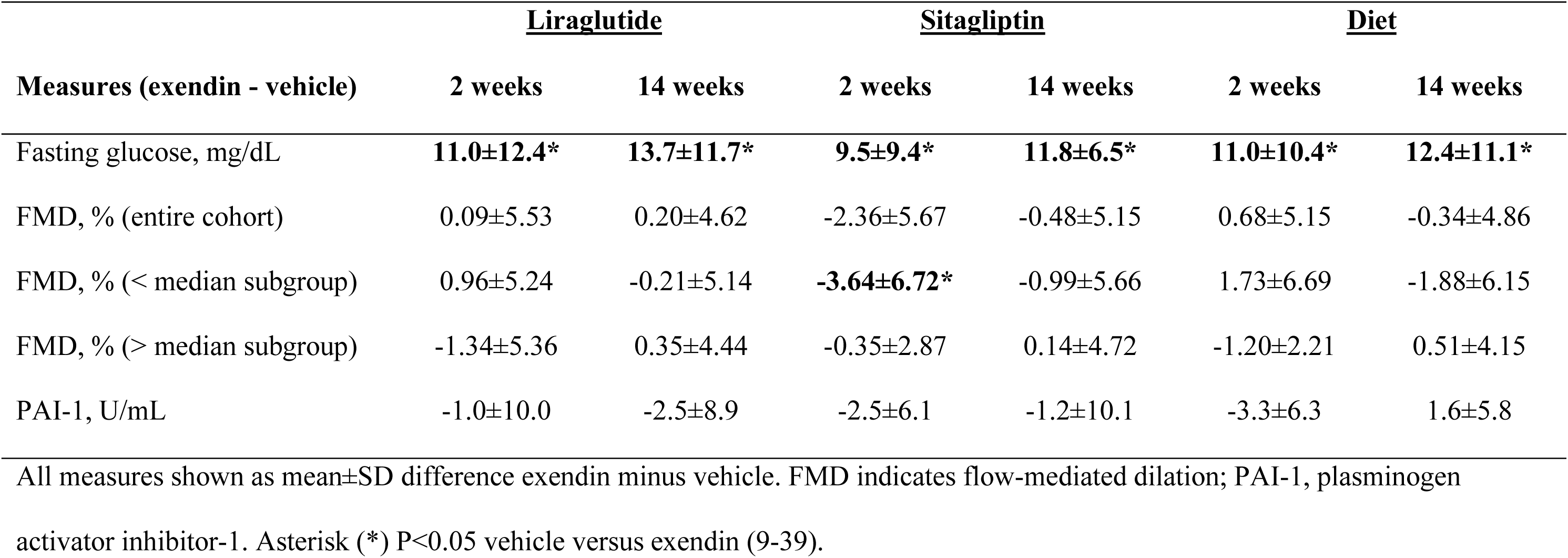
Effects of Exendin (9-39)

## Discussion

GLP1R agonists cause weight loss in obese individuals and reduce major adverse cardiovascular outcomes in T2DM. We performed a randomized controlled study comparing the effects of treatment with a GLP1R agonist, hypocaloric diet-induced weight loss, and a DPP4 inhibitor in obese pre-diabetic individuals. Our aim was to dissect the weight loss-dependent and -independent effects of GLP1R agonist treatment on measures of vascular function. We found no effect of liraglutide, diet-induced weight loss, or sitagliptin on endothelium-dependent vasodilation as measured by FMD in the overall cohort. In a post hoc analysis, liraglutide, weight loss, and sitagliptin all improved endothelial function in those individuals with reduced baseline FMD after 2 and 14 weeks. In contrast, liraglutide treatment and weight loss, but not sitagliptin treatment, significantly reduced PAI-1 after 14 weeks, and GLP1R antagonism did not alter this effect. Finally, liraglutide treatment, but neither sitagliptin nor hypocaloric diet, reduced circulating MCP-1.

Prior studies of the effects of GLP1R agonists on endothelial function as measured by FMD were all performed in individuals with T2DM and demonstrated mixed results. Two studies found no effect [18, 19], three reported improvement in FMD [20-22], and a systematic review and meta-analysis did not find a significant effect of GLP1R agonists on FMD [23]. Interpretation of these data are limited by the modest number of and heterogeneity between published studies. Our study is the largest evaluating the effect of GLP1R agonist treatment on FMD to date, and the first study that is controlled and blinded (to drug assignment). Our negative findings in the overall cohort agree with multiple prior studies and with the meta-analysis, suggesting that the cardiovascular benefits of GLP1R agonists are not mediated through improved endothelial vasodilator function as measured by FMD. The lack of an acute effect of the GLP-1R antagonist on endothelium-dependent vasodilation during liraglutide, despite dramatic effects on fasting glucose, further argues against a significant contribution of improved endothelium-dependent vasodilation to the cardiovascular protective effects of GLP1R agonists.

Notably, our overall cohort demonstrated relatively intact baseline endothelium-dependent vasodilation with a median FMD 9.91% in our group compared to values ranging from 1.6-8.9% in the prior studies [18-22]. While we anticipated higher FMD in individuals with pre-diabetes as compared to those with T2DM at the outset, prior studies had demonstrated measurable endothelial dysfunction in pre-diabetic individuals, which informed our study population choice [13]. Our exploratory analysis suggests that both GLP1R activation and weight loss improve FMD in individuals with attenuated baseline FMD, and this needs to be investigated in future studies. Our findings in the subgroup with baseline FMD above median also demonstrate that improvements beyond normal do not occur, as we have shown previously [24].

PAI-1 is the primary inhibitor of tissue plasminogen activator and is central to regulating fibrinolysis. PAI-1 is increased in obese [25] and insulin resistant individuals [26-28], and PAI-1 levels predict incident myocardial infarction [29]. PAI-1 expression is upregulated by inflammatory stimuli such as interleukin-6 and tumor necrosis factor-alpha, activation of the renin-angiotensin-aldosterone system, as well as by metabolic stimuli including hyperglycemia, hyperinsulinemia, and elevated VLDL [28]. PAI-1 decreases after weight loss [30] or treatment with GLP1R agonists [31, 32]. In this study, liraglutide reduced PAI-1 as early as 2 weeks, with concomitant reductions in blood glucose, HOMA-IR, and MCP-1, but prior to significant weight loss. In contrast, hypocaloric diet did not reduce PAI-1 at 2 weeks, despite significant reductions in weight and HOMA-IR. This suggests that the liraglutide-induced reduction in PAI-1 at 2 weeks is not driven by weight loss or improvements in insulin resistance but may be due to improvements in hyperglycemia or inflammation. The significant correlation between PAI-1 and MCP-1 levels at 2 weeks in the liraglutide group supports this notion. By 14 weeks, reductions in PAI-1 in the liraglutide and hypocaloric diet-treated groups are likely due to a combination of the above factors including additional weight loss in both groups.

MCP-1 is a key chemokine that recruits monocytes into tissues, including the vasculature, and MCP-1 levels are increased by vascular endothelial injury, oxidative stress, and inflammatory cytokines [33, 34]. MCP-1 expression is higher in diseased human arteries with atherosclerotic plaques [35, 36], and circulating MCP-1 levels predict restenosis after coronary angioplasty [37] and are associated with histopathologic markers of plaque vulnerability [38]. Furthermore, the therapeutic potential of targeting the MCP-1 pathway in cardiovascular disease has been demonstrated in animals [39]. A smaller study of liraglutide 1.2 mg versus hypocaloric diet in type 2 diabetic patients for four months reported increased circulating and subcutaneous adipose tissue MCP-1 levels with liraglutide treatment, suggesting that GLP1R agonist treatment does not improve this inflammatory parameter [40]. Yet several reports have found that activation of the GLP1R pathway reduces circulating MCP-1, although without a diet-induced weight loss comparator [41-43]. In the current study, the liraglutide-induced reduction in MCP-1 by 2 weeks, prior to clinically significant weight loss, combined with failure of hypocaloric diet-induced weight loss to lower MCP-1, suggest that activation of the GLP-1 pathway specifically lowers MCP-1 in a weight-loss independent mechanism.

The results of the current study argue against GLP-1 receptor-driven improvement in endothelium-dependent vasodilation as the basis for the beneficial cardiovascular effects seen with these drugs. Reduction of PAI-1 with liraglutide as early as 2 weeks after treatment initiation, and prior to significant weight loss, suggests a role for fibrinolysis and inflammation in the improvement. In addition, the finding that MCP-1 was uniquely reduced during treatment with liraglutide may provide insight into the mechanism of cardiovascular benefit with GLP1R agonists. Another mechanism of cardiovascular benefit may involve platelet activation. Platelet activation plays a critical role in atherosclerosis and thrombosis, and platelets express GLP1R. Barale et al have shown that GLP1R agonists potentiate the anti-aggregating effects of nitric oxide in human platelets [44]. We have recently reported that liraglutide attenuates thromboxane-induced platelet aggregation both *in vitro* and *in vivo* (Cahill KN et al. JACC: Basic to Translational Science, *manuscript in press*). Of note, MCP-1 expression is increased by platelet-derived growth factor [45], and activated platelets induce MCP-1 release *in vitro* [46]. While we did not detect an effect of any treatment on circulating P-selectin, P-selectin reflects both platelet and endothelial cell activation [33]. Thus, further studies are needed to assess the antithrombotic effects of GLP1R agonists.

Weight loss in the hypocaloric diet group was associated with a significant decline in systolic blood pressure over 14 weeks, whereas there was no decrease in systolic blood pressure in either the sitagliptin or liraglutide group. Weight loss was significantly less in the liraglutide-treated group compared to the hypocaloric diet-treated, and GLP1R agonist-mediated stimulation of the sympathetic nervous system may have counteracted any favorable effect of weight loss on systolic blood pressure in the liraglutide-treated group. Activation of GLP-1 receptors in the hypothalamus and brainstem by long-acting GLP1R agonists stimulate sympathetic outflow [47]. Increased heart rate, observed in the liraglutide-treated group, is a hallmark of treatment with stable GLP1R agonists [48]. Interestingly, treatment with sitagliptin did not increase heart rate. We have previously reported that heart rate is increased after short-term treatment with combined sitagliptin and ACE inhibition [49, 50], but not after longer duration treatment [51], even though norepinephrine concentrations are increased. Decreased degradation of substance P may contribute to sympathetic stimulation when both DPP4 and ACE are inhibited. In the present study, a minority of participants were taking concurrent anti-hypertensive medications.

This study has several limitations. The majority of participants enrolled were women and the study was not powered to permit comparison between effects in men and women. In addition, we enrolled individuals with pre-diabetes rather than overt diabetes, which may have diminished the effect size of our treatment by eliminating the hyperglycemia component of vascular dysfunction [52]. We chose to enroll obese pre-diabetic individuals to study a homogenous population with baseline insulin resistance but without overt hyperglycemia to avoid confounding by concurrent anti-diabetic medications or by varying severity and duration of diabetes. As noted, the participants had normal baseline endothelium-dependent vasodilation. We completed a post hoc analysis stratified by baseline FMD below and above the median. Although not pre-specified, the finding that those with lower baseline FMD were significantly more insulin resistant than those with higher baseline FMD provides physiologic support of the analysis. However, all three interventions improved FMD in this subgroup to some extent, and clinical data has proven that only liraglutide, and not sitagliptin or equivalent weight loss through diet, improve cardiovascular outcomes.

In conclusion, in the largest study of the effect of a GLP1R agonist on endothelium-dependent vasodilation, liraglutide, sitagliptin, and diet-induced weight loss had no effect on FMD in obese pre-diabetic individuals with normal baseline endothelial function. Liraglutide, sitagliptin and diet all improved FMD in those individuals with baseline endothelial dysfunction. Liraglutide appears to have anti-inflammatory (MCP-1 and PAI-1) and anti-thrombotic (PAI-1) effects that precede weight loss. Reduction of MCP-1 by liraglutide differentiates the effects of the GLP1R agonist from weight loss or DPP4 inhibition. Such an anti-inflammatory effect could contribute to the unique cardiovascular benefit of GLP1R agonists.

## Data Availability

All data produced in the present study are available upon reasonable request to the authors.

## Abbreviations

DPP4: Dipeptidyl peptidase 4
FMD: Flow-mediated dilation
GLP-1: Glucagon-like peptide-1
GLP1R: Glucagon-like peptide-1 receptor
HbA1c: Hemoglobin A1c
HOMA-IR: Homeostatic Model Assessment for Insulin Resistance
HOMA2: Homeostatic Model Assessment 2
MCP-1: monocyte chemoattractant protein-1
NMD: Nitroglycerin-mediated dilation
OGTT: Oral glucose tolerance test
PAI-1: Plasminogen activator inhibitor-1
T2DM: Type 2 diabetes mellitus
UACR: Urine albumin-to-creatinine ratio

## Acknowledgements

The authors acknowledge contributions from Anthony Dematteo.

## Data Availability

The datasets generated during and/or analyzed during the current study are available from the corresponding author on reasonable request.

## Authors’ Relationships and Activities

None unless noted below:

J.A.B.: Dr. Beckman is a consultant for JanOne, serves on a DSMB for Janssen and Novartis, and has ownership in EMX and Janacare.

J.R.K: Dr. Koethe has served as a consultant to Gilead Sciences, Merck, ViiV Healthcare, Theratechnologies and Janssen. He has also received research support from Gilead Sciences and Merck.

J.M.L: Dr. Luther has served on the advisory board for Mineralys.

N.J.B.: Dr. Brown serves on the scientific advisory board for Alnylam Pharmaceuticals. She serves as a consultant for Pharvaris Gmbh and eBioStar Tech. Dr. Brown owns equity in Abbvie and J and J Pharmaceuticals.

## Contribution Statement

M.M. researched data and wrote the manuscript. J.A.B. contributed to study design and edited the manuscript. H.N. performed statistical analysis. E.M.G. researched data. D.M. designed the database and managed study days. P.W. managed study days. S.E.H. researched data. B.P. researched data. D.O. researched data. W.E.S. researched data. J.K.D. contributed to study design. J.R.K. contributed to study design and edited the manuscript. J.D.B. contributed to study design and edited the manuscript. K.N.C. researched data. C.Y. performed statistical analysis. H.S. designed and managed the diet intervention and assessments, researched data, and edited the manuscript. K.N. contributed to study design, obtained funding and edited the manuscript. J.M.L. researched data and edited the manuscript. N.J.B. designed the study, obtained funding, researched data, managed the team, and edited the manuscript.

Guarantor for the study is Nancy J. Brown.

