## Supplemental Materials for "Comparative Effects of Weight Loss and Incretin-Based Therapies on Vascular Endothelial Function, Fibrinolysis, and Inflammation: A Randomized, Controlled Trial"

**Electronic Supplementary Material**

**Detailed Methods**

Full trial protocol can be obtained by e-mailing the corresponding author and is also available on clinicaltrials.gov NCT03101930.

Medication management in run-in phase

Lipids were managed using the current American Heart Association guidelines. Individuals who smoked received smoking cessation recommendations. Hypertensive individuals were treated to target blood pressure goal of 140/90 mmHg with a standardized regimen. This included prescription or optimization of prescription of angiotensin-converting enzyme (ACE) inhibitor or angiotensin receptor blocker (ARB), followed by the addition of amlodipine was recommended as a second agent, followed by chlorthalidone. Finally, participants were advised on the use of low dose aspirin 81mg based on US Preventative Services Task Force guidelines.

Vascular endothelial measures

Measurements were made as previously reported.[1-4] Brachial artery diameter was measured using B-mode ultrasonography equipped with a high resolution linear array transducer (7.5 mHz). A longitudinal image (parallel to the artery) was acquired just proximal to the antecubital fossa with the transducer positioned to optimize images of the near and far wall interfaces. A simultaneous electrocardiographic signal was recorded, and images are digitally acquired at end-diastole, synchronized to the R wave on the electrocardiogram. To assess endothelium-dependent vasodilation, brachial artery diameter was measured under basal conditions and during reactive hyperemia. Reactive hyperemia results after five min of ischemia produced by inflating a BP cuff on the upper arm to suprasystolic pressures. We have found that following cuff deflation, the maximal increase in brachial artery diameter occurs at approximately one min of reactive hyperemia, a response mediated by endothelium-derived nitric oxide.[2] After a rest period, endothelium-independent vasodilation was assessed by imaging the brachial artery under basal conditions and following the administration of sublingual nitroglycerin (0.4 mg). Maximal brachial artery dilation occurs three to four min after sublingual nitroglycerin. The video output and electrocardiographic signal of the ultrasound machine was connected to a computer equipped with a Data Translation frame grabber videocard. The R wave on the electrocardiogram was used as a trigger to acquire frames. Digitized images was stored on the hard drive and backed up on removable media. Acquisition and analysis of the stored images was performed using software designed for this purpose by Medical Imaging Applications. The vessel wall lumen interface was determined by derivative-based edge detection following identification of the region of the anterior and posterior walls by the investigator. The maximum diameter of the vessel was determined and the percent change in diameter calculated.

Homeostatic Model Assessment of Insulin Resistance (HOMA-IR)

HOMA-IR was calculated from the fasting measurements using the following formula: (insulin (µU/mL)*glucose(mg/dL))/405.

HOMA2

HOMA2 was calculated using the following calculator: https://www.dtu.ox.ac.uk/homacalculator/.

**Supplementary Figure S1**

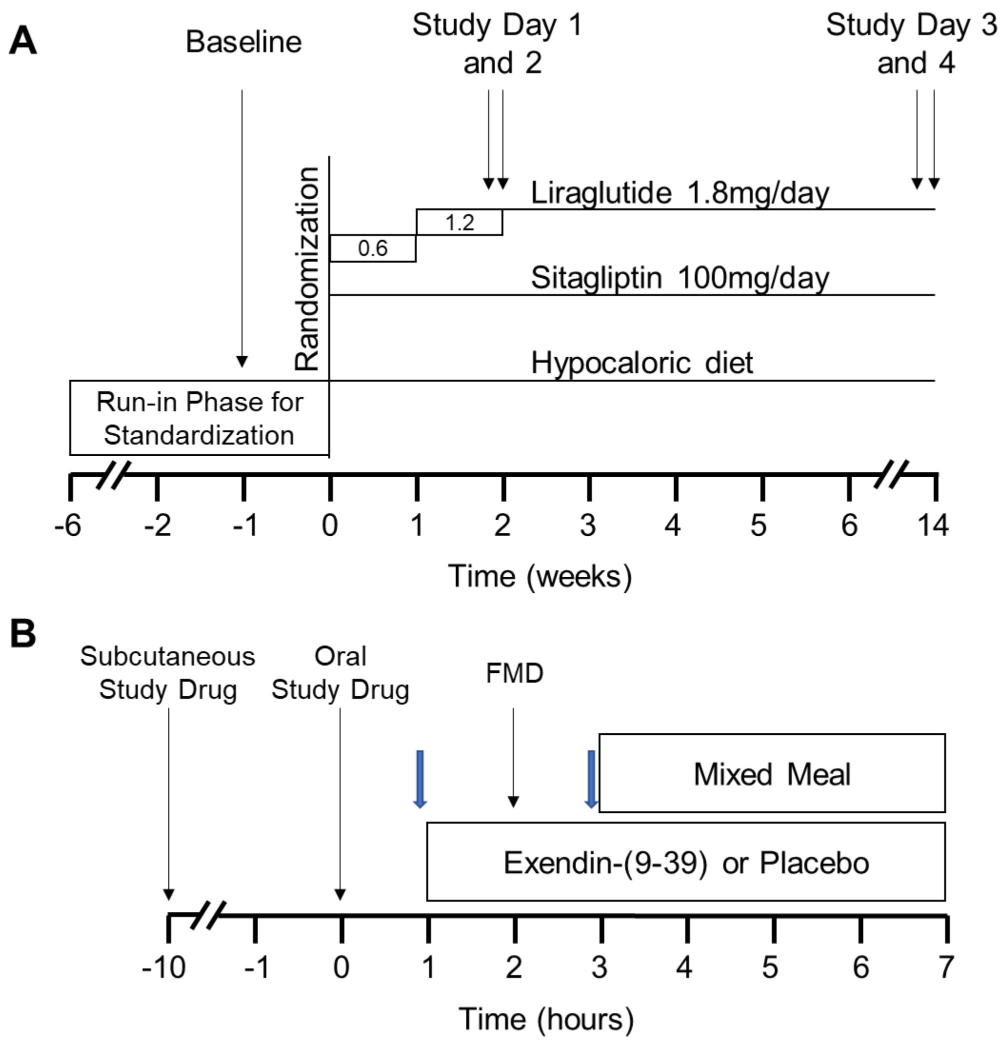


**Supplemental Figure 1. Study Protocol. (A)** Timeline of the overall study protocol including run-in phase, randomization, baseline visit and study days. **(B)** Details of the study day protocol. Blue block arrows indicate timing of pre- and post-infusion blood collection. FMD indicates flow-mediated dilation.

**Supplementary Figure S2**

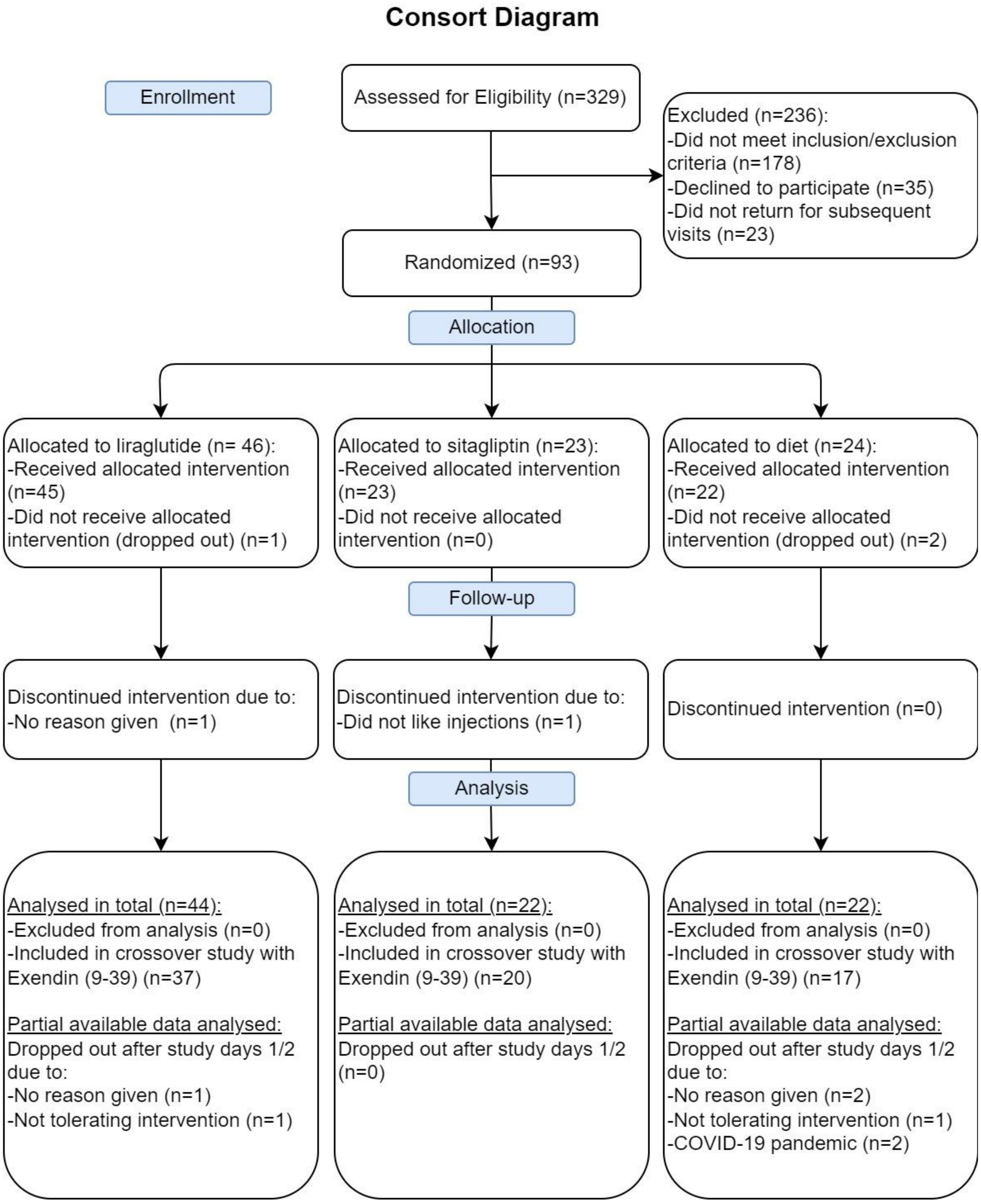

**Supplementary Figure S3**

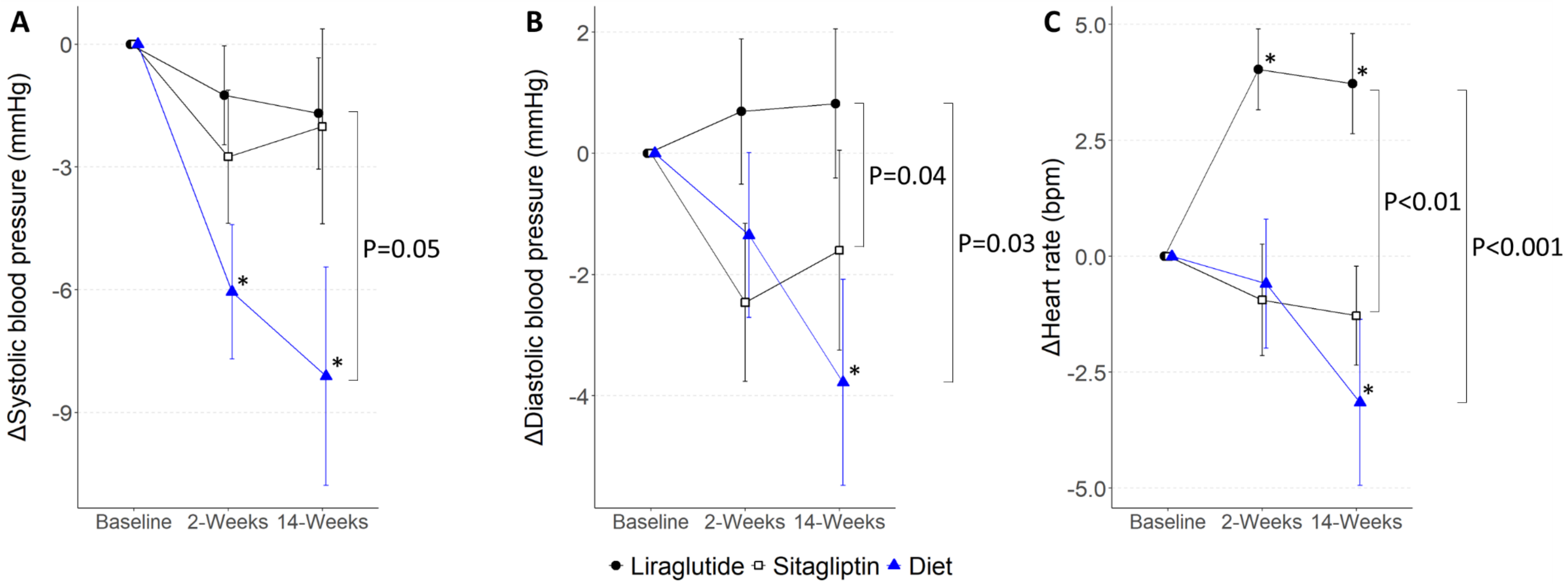


**Supplemental Figure 3. The Effect of Treatment on Hemodynamic Measures.** Plots show mean ± SEM for **(A)** systolic blood pressure, **(B)** diastolic blood pressure, and **(C)** heart rate at 2 and 14 weeks of treatment as change from baseline. Asterisk (*) symbols indicate P<0.05 for estimates of change from baseline, and brackets indicate difference between treatments at 14 weeks.
